# Procurement of innovation terminology usage in health care: A scoping review protocol

**DOI:** 10.1101/2020.01.17.19015107

**Authors:** Margaret R. Andrews, Romualdo Ramos, Martina Ahlberg, Jan A. Hazelzet, Erik M. van Raaij, Vikrant Sihag, Rossana Alessandrello, Eva Aurín, Bahar Bakir, Desmond Carter, Suzan Ikävalko, Mariet Nouri Janian, Elettra Oleari, Maarten M. Timmermann, Victòria Valls-Comamala, Tanja A. Stamm, Platform for Innovation of Procurement and Procurement of Innovation Project

## Abstract

**Background:** Although procurement of innovation is an established policy tool used to stimulate collaboration between supply- and demand-side entities during the development of new technologies, there is little scientific literature describing the process as applied in health care settings. Furthermore, what literature exists contains inconsistencies of terms, definitions, and/or concepts related to procurement of innovation. This protocol details our process for a systematic scoping review to describe the current scope of literature and to provide terminology clarification.

**Methods:** A search strategy will be used to search PubMed, EMBASE [OVID], CINAHL [EBSCO], PsycINFO [ProQUEST], ABI/INFORM, ISI Web of Knowledge, EBSCO, JSTOR, the Cochrane Database of Systematic Reviews, and Google Scholar; grey literature, non-scientific reports, policy documents and expert recommendations will also be considered as additional sources for texts. Two researchers will screen titles and abstracts for inclusion/exclusion criteria, followed by full texts. We will extract the following data, if applicable: title, authors, date, author affiliations, country, journal/publication characteristics, setting, aims/purpose, methodology, sample characteristics, assessment/evaluation tools, outcome parameters, key findings, relevance, and terminology usage/definitions. Results will be presented narratively and visually.

**Discussion:** This paper describes the steps of our proposed systematic scoping review to identify and analyse scientific and non-scientific literature related to procurement of innovation and/or innovation of procurement in health care settings, with a particular focus on digital health technologies. Results are intended to demonstrate the current scope of literature, to provide clarity in language and therefore to serve as a first step for further research in this growing field.

## Introduction

Medical technologies of various types are used in ever increasing ways by providers and consumers across the entire spectrum of health care. New technologies offer great opportunities for health care systems, however they can also represent significant portions of health care costs. For this reason, it is a priority for many health care managers in strategic leadership positions to transition to the usage and development of medical technologies that balance cost and utility (1, 2). One way to do this is to ensure that the diffusion and uptake of new technologies is maximised (3), which can be positively impacted by the degree of involvement of end-users in the purchasing and/or development of new products (4, 5).

The development and usage of new digital health solutions is a prime example of this phenomenon. Rapid expansion of products and services has been driven from all sides, but the supply side has primarily controlled new development (6, 7). It is not uncommon that the subsequent uptake and diffusion of these products is suboptimal (3, 7, 8). One potential reason for this is a disconnect between the real needs of health care stakeholders and the solutions designed by digital technology developers (9). A number of efforts to enhance collaboration between the supply and demand sides have therefore been fostered; among these is the European Union’s (EU) stimulation of the procurement of innovation in health care.

In 2018, the EU issued a new Guidance on Innovation Procurement (10) document that built upon the 2014 modernised public procurement directives. These publications were part of the EU’s efforts to stimulate innovative research and development (R&D) through increased support of public procurement actions in different sectors. Such efforts have not been restricted to Europe; indeed, procurement of innovation is used globally to foster R&D (2–4, 11).

To-date, however, information on these tools and their usage in health care settings has primarily been published in policy documents and not widely discussed in scientific literature (11–13). Furthermore, literature contains differing terminology, definitions, and/or concepts related to procurement of innovation (1, 3, 9, 12, 14, 15). In comparison, some of these terms have specific definitions in each policy document. In the EU’s 2018 Guidance (10), (public) innovation procurement is loosely defined as all procurement actions that either: a) purchase the process of innovation (i.e. research and development) and resulting full or partial outcomes; or, b) purchase the outcome(s) of innovation from another party. However, we are currently lacking single definitions of these terms which are agreed upon and used identically by experts. Differences in terminology decrease the possibility to compare effectively between documents, policies and projects, and represent a serious barrier to a common understanding in the respective field(s).

We therefore propose to perform a scoping review of existing scientific and grey literature in order to provide clarification of terminology. More scientific research is needed to build a body of evidence on the effects of procurement of innovation, particularly in health care. Terminology clarification is a first step in supporting this effort.

### Objectives

The objective of this research is to clarify and better understand terminology related to procurement of innovation and innovation of procurement. A scoping review will be performed. Relevant information and results will be extracted and applied to health care innovations in general, and to the area of digital solutions in particular. Finally, we will consider how the procurement of innovation fits into broader developments in health care.

### Research Questions

This research will seek to answer the following questions within the context of health care and especially in relation to digital health:

1. What are the relevant definitions of “procurement of innovation” and “innovation of procurement” and/or how have the terms been used to-date?
2. What are the essential elements which characterise the terms related to procurement of innovation and innovation of procurement? E.g., how is procurement of innovation different from more traditional (public) procurement?
3. How does the procurement of innovation fit into broader developments in the field?

### Keywords

Purchasing, procurement, public procurement, procurement of innovation, innovation of procurement, pre-commercial procurement, public procurement of innovation; innovation, medical technology, digital health; health care, hospital; scoping review

## Methods

This review will be completed using the Joanna Briggs Institute’s (JBI) approach to evidence-based healthcare methodology (16).

### Inclusion Criteria

#### Types of studies

The following types of publications will be eligible for inclusion: policy papers and opinion pieces; quantitative, qualitative, intervention, mixed-methods, and case studies; literature reviews of all types; expert opinions; conference proceedings; book chapters; official reports/documents; and relevant websites, especially governmental sites.

Studies/articles/reports will be excluded if they are not written in English, German, Dutch, Spanish, Swedish, Italian, Finnish, or French.

#### Problem/Participants

If applicable, any type of participant, whether patients/consumers of health care or health care providers, will be considered for this review. This may include populations of individuals, health care providers at the institution level, and/or health care systems at the local, state, national, or international levels.

#### Concept

Publications will be included in this review if they discuss procurement (of technology) and/or innovation in a health care setting.

This review will include publications concerning any type of intervention, as well as publications where no intervention took place such as policy documents, reports and expert opinions. However, we are specifically looking for the following information:

- Interventions or case studies where a health care provider was involved with the development of new products, designs, methods, or services relating to medical technologies;
- Interventions or case studies where new products, designs, methods, or services relating to medical technologies were developed, implemented, and/or evaluated;
- Interventions or case studies where the procurement of new products, designs, methods, or services relating to medical technologies in health care settings was stimulated, tested, or evaluated;
- Publications discussing new products, designs, methods, or services relating to medical technologies;
- Publications discussing procurement of new products, designs, methods, or services relating to medical technologies.

#### Context

Health care settings of all types.

### Search strategy

The search will be completed in three steps. An initial limited search will be conducted to verify our selection of search terms and current proposed strategy (see Table I and Appendix I). If needed, the text words and index terms may be modified based on an analysis of the results of this initial search. In the second step, a full query will be performed in the following databases: PubMed, EMBASE [OVID], CINAHL [EBSCO], PsycINFO [ProQUEST], ABI/INFORM, ISI Web of Knowledge, EBSCO, JSTOR, the Cochrane Database of Systematic Reviews, and Google Scholar. The search for unpublished studies and other types of publications will include: OpenGrey – System for Information on Grey Literature in Europe; University of York Centre for Reviews and Dissemination; Library Hub Discover; national and international websites and databases related to the topic. Task-force members will be consulted to identify additional grey literature or research that has not been found through the database query. In the third step, we will search the reference lists of identified papers for any additional relevant articles, which will be subjected to the same screening and selection process. We will limit the search to titles and abstracts; however, we will not limit the search to a specific date range.

**Table I:** search terms

| Concept: Procurement | Concept: Innovation | Context |
| --- | --- | --- |
| Procure, procurement, procuring, purchase, purchasing, source, sourcing, tender, tendering<br>"Group Purchasing" [Mesh]<br>"Purchasing, Hospital" [Mesh]<br>"Value-Based Purchasing" [Mesh] | Innovate, innovation, innovative, technology, technological, device<br>new software development, new hardware development, new product development, telemedicine, digital health, digital solution, mHealth, eHealth, mobile health, electronic health | Health care, healthcare, medical, hospital, clinic, clinical<br>"Health Occupations" [Mesh]<br>"Health Care Category" [Mesh] |
| pre-commercial procurement, procurement of innovation, innovation procurement, innovative procurement, medical device purchasing, medical device procurement |  |  |

## Study selection

As an initial step, identified publications will be uploaded into EndNote and duplicates removed. The selection process will have two phases. In the first phase, two researchers (MRA, RR) will independently assess all English- and German-language titles and abstracts to determine whether the inclusion criteria have been met. Task Force members will review titles and abstracts of items in Dutch, Spanish, Swedish, Italian, Finnish, and French. In the second phase, full texts for all selected publications will be analysed. Any discrepancies between reviewers and/or questions as to whether a publication has met the inclusion criteria will be resolved by a consensus decision with an expert (TAS) and/or the Task Force. These results, including interrater agreement and reasons for exclusion, will be presented in a PRISMA flow diagram in the subsequent review (Appendix II). Reasons for exclusion of full-text publications will furthermore be provided in accordance with the PRISMA Extension for Scoping Reviews (PRISMA-ScR) statement (17, 18) in an appendix.

## Data extraction

Data will be extracted using a standardized extraction form based on the methodology for scoping reviews developed by the Joanna Briggs Institute (16). We will extract the following data, if applicable: title, authors, date, author affiliations, country, journal/publication characteristics, setting, aims/purpose, methodology, sample characteristics, assessment/evaluation tools, outcome parameters, key findings, relevance, and terminology usage/definitions (Appendix III). As necessary, this extraction form may be modified throughout the review process; any revisions will be detailed in the full review. Two reviewers (MRA, RR) will extract data independently. Any disagreements that arise between the reviewers will be resolved through discussion with an expert (TAS) and/or the Task Force, and a consensus decision reached.

## Assessment of risk of bias

According to the established standards for scoping reviews, no formal assessment of methodological quality or risk of bias will be conducted for the selected publications (16, 18).

## Data mapping

According to the objectives of this review, results will be presented in a variety of ways including narratively and visually. Definitions will be presented in a table. Further characteristics may be presented in tabular, diagrammatic/graphic, or logical format. The presentation of results will be refined and finalized during the review.

## Data Availability

Data availability is not relevant at this time because the submission is a scoping review protocol and no data exists. Extracted data in the final publication will be attached in an appendix and/or available upon request.

## Appendix I: Search strategy for PubMed

**1) Population: not applicable**

**2) Concept: procurement, innovation**

(((((((((((procure[Title/Abstract]) OR procurement[Title/Abstract]) OR procuring[Title/Abstract]) OR purchasing[Title/Abstract]) OR purchase[Title/Abstract]) OR source[Title/Abstract]) OR sourcing[Title/Abstract]) OR tender[Title/Abstract]) OR tendering[Title/Abstract]) OR “Group Purchasing” [Mesh]) OR “Purchasing, Hospital” [Mesh]) OR “Value-Based Purchasing” [Mesh]) AND

(((((((((((((((innovate[Title/Abstract]) OR innovation[Title/Abstract]) OR innovative[Title/Abstract]) OR technology[Title/Abstract]) OR technological[Title/Abstract]) OR device[Title/Abstract]) OR new software development[Title/Abstract]) OR new hardware development[Title/Abstract]) OR new product development[Title/Abstract]) OR telemedicine[Title/Abstract]) OR digital health[Title/Abstract]) OR digital solution[Title/Abstract]) OR mHealth[Title/Abstract]) OR eHealth[Title/Abstract]) OR mobile health[Title/Abstract]) OR electronic health[Title/Abstract]) AND

**3) Context: health care setting**

(((((((health care[Title/Abstract]) OR healthcare[Title/Abstract]) OR medical[Title/Abstract]) OR hospital[Title/Abstract]) OR clinic[Title/Abstract]) OR clinical[Title/Abstract]) OR “Health Occupations” [Mesh]) OR “Health Care Category” [Mesh])

**4) Combined phrases**

((((((pre-commercial procurement[Title/Abstract]) OR procurement of innovation[Title/Abstract]) OR innovation procurement[Title/Abstract]) OR innovative procurement[Title/Abstract]) OR medical device purchasing[Title/Abstract]) OR medical device purchasing[Title/Abstract])

## Appendix II: PRISMA flow diagram (17)

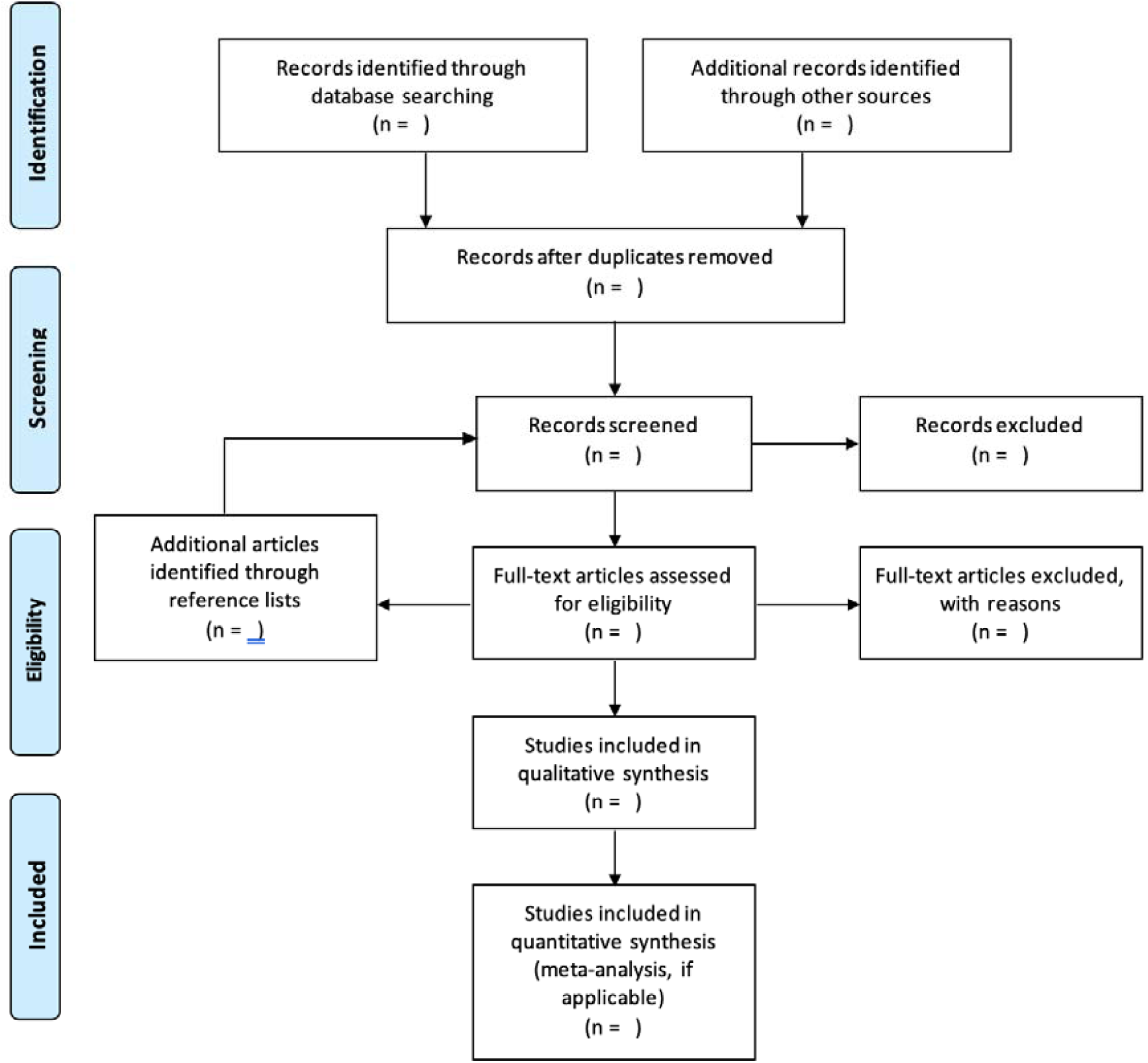

## Appendix III: Extraction Form

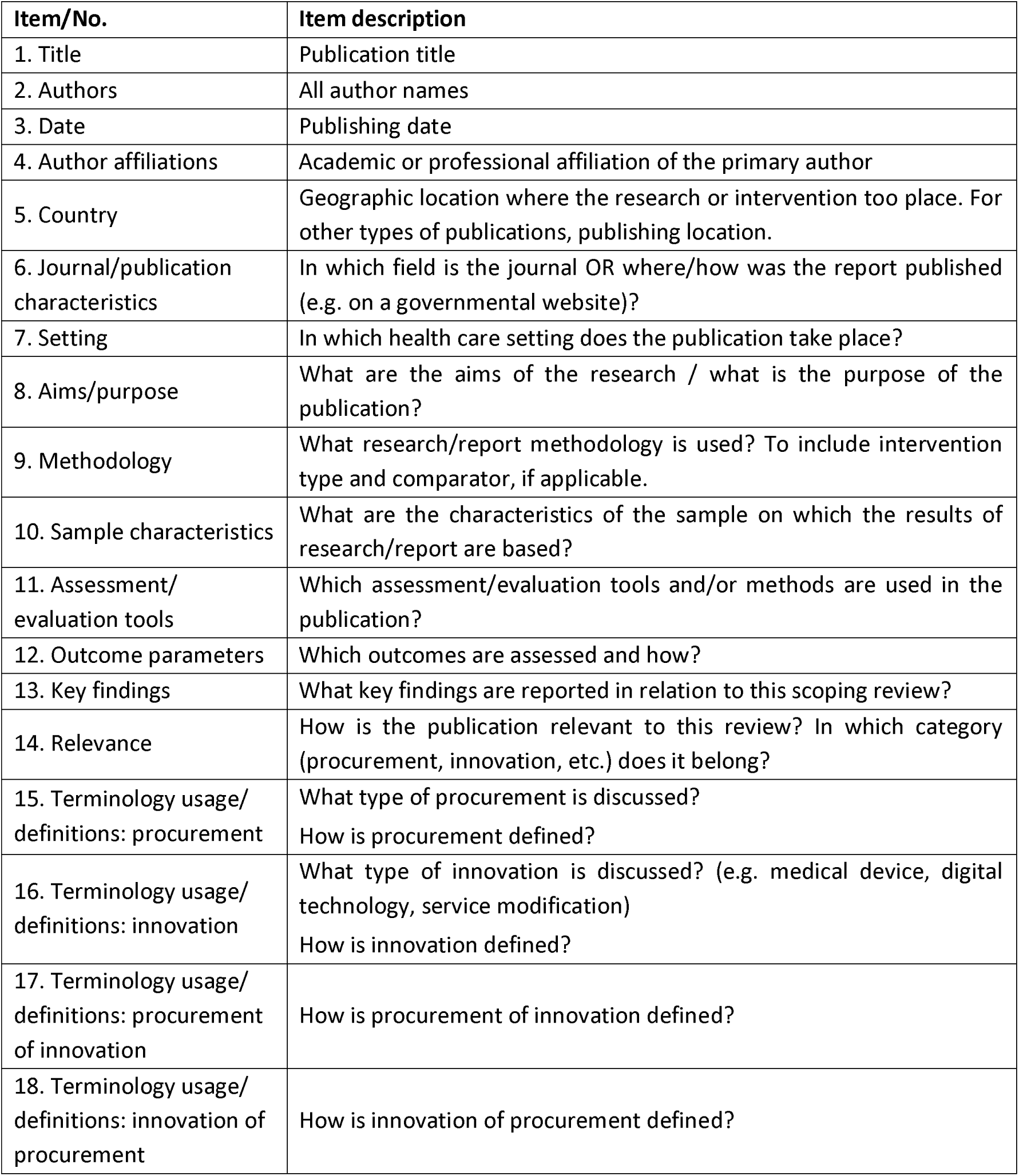

| Item/No. | Item description |
| --- | --- |
| 1. Title | Publication title |
| 2. Authors | All author names |
| 3. Date | Publishing date |
| 4. Author affiliations | Academic or professional affiliation of the primary author |
| 5. Country | Geographic location where the research or intervention took place. For other types of publications, publishing location. |
| 6. Journal/publication characteristics | In which field is the journal OR where/how was the report published (e.g. on a governmental website)? |
| 7. Setting | In which health care setting does the publication take place? |
| 8. Aims/purpose | What are the aims of the research / what is the purpose of the publication? |
| 9. Methodology | What research/report methodology is used? To include intervention type and comparator, if applicable. |
| 10. Sample characteristics | What are the characteristics of the sample on which the results of research/report are based? |
| 11. Assessment/evaluation tools | Which assessment/evaluation tools and/or methods are used in the publication? |
| 12. Outcome parameters | Which outcomes are assessed and how? |
| 13. Key findings | What key findings are reported in relation to this scoping review? |
| 14. Relevance | How is the publication relevant to this review? In which category (procurement, innovation, etc.) does it belong? |
| 15. Terminology usage/definitions: procurement | What type of procurement is discussed?<br>How is procurement defined? |
| 16. Terminology usage/definitions: innovation | What type of innovation is discussed? (e.g. medical device, digital technology, service modification)<br>How is innovation defined? |
| 17. Terminology usage/definitions: procurement of innovation | How is procurement of innovation defined? |
| 18. Terminology usage/definitions: innovation of procurement | How is innovation of procurement defined? |

